# A Single Dose of COVID-19 mRNA Vaccine Induces Airway Immunity in COVID-19 Convalescent Patients

**DOI:** 10.1101/2021.12.16.21267932

**Authors:** Emanuela Martinuzzi, Jonathan Benzaquen, Olivier Guerin, Sylvie Leroy, Thomas Simon, Marius Ilie, Véronique Hofman, Maryline Allegra, Virginie Tanga, Emeline Michel, Jacques Boutros, Charlotte Maniel, Antoine Sicard, Nicolas Glaichenhaus, Cecil Czerkinsky, Philippe Blancou, Paul Hofman, Charles Marquette

## Abstract

**Background:** Mucosal antibodies can prevent virus entry and replication in mucosal epithelial cells and hence virus shedding. Preclinical and clinical studies have shown that a parenteral booster injection of a vaccine against a mucosal pathogen promotes stronger mucosal immune responses following prior infection compared to two injections of a parenteral vaccine. We investigated whether this was also the case for a COVID-19 mRNA vaccine.

**Methods:** Twenty-three COVID-19 convalescent patients and 20 SARS-CoV-2-naive subjects were vaccinated with respectively one and two doses of the Pfizer-BioNTech COVID-19 RNA vaccine. Nasal Epithelial Lining Fluid (NELF) and plasma were collected before and after vaccination and assessed for Immunoglobulin (Ig)G and IgA to Spike and for their ability to inhibit the binding of Spike to its ACE-2 receptor. Blood was analyzed one week after vaccination for the number of Spike-specific Antibody Secreting Cells (ASCs) with a mucosal tropism.

**Results:** In COVID-19 convalescent patients, a single dose of vaccine amplified pre-existing Spike-specific IgG and IgA antibody responses in both NELF and blood against both vaccine homologous and variant strains, including delta. These responses were associated with Spike-specific IgG and IgA ASCs with a mucosal tropism in blood. Nasal IgA and IgG antibody responses were lower in magnitude in SARS-CoV-2-naive subjects after two vaccine doses

**Conclusion:** This study showed that a parenteral booster injection of a COVID-19 RNA vaccine promoted stronger mucosal immune responses in COVID-19 convalescent patients compared to SARS-CoV-2 naive subjects who had received a first vaccine dose.

## Introduction

Amid the surge of variants of concern, most COVID-19 vaccines are highly effective against hospitalization and death of patients caused by a variety of SARS-CoV-2 strains (1) (2-4). Early studies performed before the emergence of the delta variant showed that individuals who were fully vaccinated with an mRNA vaccine were less likely than unvaccinated persons to be infected with SARS-CoV-2 or to transmit it to others (5, 6). Likewise, with the delta variant, studies have shown that vaccination reduces the risk of infection. Nonetheless, fully vaccinated individuals with breakthrough infections still transmit infection, including to fully vaccinated contacts (7). Therefore, the risk of a SARS-CoV-2 breakthrough infection in fully vaccinated people cannot be eliminated as long as there is continued transmission of the virus.

Protection from severe forms of COVID-19 in fully vaccinated adults is mediated, at least in part, by SARS-CoV-2-specific antibodies, as demonstrated by the transfer of plasma from COVID-19 convalescent individuals to recently infected patients (8). However, the immune mechanisms that prevent carriage and shedding of SARS-CoV-2 remain to be elucidated. Lessons drawn from other mucosal pathogens suggest that mucosal antibodies and especially secretory immunoglobulin A (sIgA) can efficiently block transmission of respiratory viruses (9). Relevant to this issue, SARS-CoV-2-specific humoral responses are dominated by IgA and peripheral expansion of IgA ASCs with a mucosal homing potential occurs shortly after disease onset (10). Further, sIgA are substantially more potent than bone marrow-derived serum monomeric IgA and IgG at neutralizing SARS-CoV-2 (11). As for Spike-specific IgA induced by mRNA COVID-19 vaccines, these antibodies are detected in SARS-CoV-2-naive individuals in plasma (10), milk (12), saliva (13) and nasal fluids (14) as early as two weeks after vaccination and for up to 6 months.

A dominant concept in vaccinology is that mucosal immunity is more efficiently induced by mucosal (ex. nasal or oral) administration of vaccines than by parenteral injection and that mucosal immune memory wanes more rapidly than systemic immune memory (12, 13). On a related topic, we and others have shown that a single booster injection of inactivated poliovirus vaccine boosted mucosal immune responses and reduced virus shedding in individuals previously vaccinated with an oral live polio vaccine (14). Building on these results, we hypothesized that immunization by parenteral injection with a COVID-19 mRNA vaccine might be more effective at inducing airway mucosal immunity in COVID-19 convalescent patients compared to SARS-CoV-2-naive subjects.

## Methods

### Study design and participants

Forty-three otherwise healthy subjects (23 SARS-CoV-2 naive and 20 COVID-19 convalescent patients) were included in a prospective monocentric longitudinal study between April 14, 2021 and June 15, 2021 at the Nice University Hospital (Nice, France). The participants who recovered from a SARS-CoV-2 infection had a history of mild or moderate COVID-19 three months or more before inclusion. These subjects will be referred thereafter as Covid^+^ while the others will be referred as Covid^-^. While all subjects received one injection of the BNT162b2 mRNA COVID-19 vaccine (Pfizer-BioNTech) on day 0, only Covid^-^ subjects received a second injection of the BNT162b2 mRNA COVID-19 vaccine on day 21, as recommended by the French National Health Authority (15). Blood was collected on day 0, 7 and 21 from all subjects, and NELF were collected on day 0 and 21 from all subjects and on day 28 and 42 for Covid-subjects (Supplementary Figure 1). All samples were centralized and stored in our Biobank (16). All subjects signed an informed consent to participate in this work. The study was approved by the CPP Sud Méditerranée V ethics committee. ClinicalTrial.gov identifier: NCT04418206.

### Nasal epithelial lining fluid (NELF)

Hydroxylated polyvinyl acetate (PVA) sponges (Merocel® Standard Dressing, ref 400400, Medtronic, Minneapolis, MN, US), were inserted between the nasal septum and the inferior turbinate (17), left in place for 3 to 6 minutes until they swelled, gently retrieved and placed in a 50 ml Falcon tube (Dustcher, Bernolsheim, France) containing 2 ml of saline solution. The fluid contained in the sponge (saline + nasal secretions) was then extracted by simple pressure, aliquoted and frozen at -70°C.

### SARS-CoV-2-specific IgG and IgA

IgA and IgG levels reacting to the SARS-CoV-2 Spike protein, the SARS-CoV-2 Spike N terminal Domain (NTD), the SARS-CoV-2 Spike Receptor Binding Domain (RBD) and the SARS-CoV-2 N protein were measured using the V-PLEX® SARS-CoV-2 Panel 2 (IgA) kit and the V-PLEX® SARS-CoV-2 Panel 2 (IgG) kit (MSD, Maryland, US), respectively. Total IgA and IgG levels were measured using the V-PLEX® Isotyping Panel 1 Human/NHP Kit total human IgA kit and the V-PLEX® total human IgG kit, respectively. Plasma and nasal fluids were diluted 100-fold and 10-fold respectively before being assessed for IgA and IgG levels. Data were acquired on the V-PLEX® Sector Imager 2400 plate reader and analysed using Discovery Workbench 3·0 software. Serial 4-fold dilutions of the standards were run to generate a 7-standard curve, and the diluent alone was used as a blank. For the plasma and NELF levels of IgA and IgG to the SARS-CoV-2 Spike full-length, NTD and RBD and N protein, the standards consisted of three levels of serum-based controls and IgA and IgG levels were expressed in arbitrary units per mL (AU/mL). For total IgA and IgG, the standards consisted of purified human IgA and IgG, and total IgA and IgG and were measured in pg/mL.

### Binding inhibition assay

Plasma and nasal fluids were assessed for antibodies inhibiting the binding of a soluble angiotensin-converting enzyme 2 (ACE2) receptor to the SARS-CoV-2 RBD derived from the Wuhan strain and from its B.1.1.7 (alpha), B.1.351 (beta), B.1.526.1 (New York), B.1.617.1 (kappa), B.1.617.2 (delta), P.1 (gamma) and P.2 (zeta) variants using the multiplex V-PLEX® SARS-CoV-2 Panel 13 ACE2 Kit. Plasma and NELF were diluted 100- and 10-fold respectively before being assessed for IgA and IgG. Data were acquired on the V-PLEX® Sector Imager 2400 plate reader and analyzed using the Discovery Workbench 3·0 software (MSD). Standard curves were generated using standards provided in the kit. Serial 4-fold dilutions of the standards were run to generate a 7-standard concentration set, and the diluent alone was used as a blank. The percentage inhibition was calculated according to the manufacturer’s instructions. This assay has been shown to correlate with assays for viral neutralization.

### EliSpot assays

To test whether the nasal mucosal IgA responses differed between COVID-19 convalescent subjects and SARS-CoV-2-naive subjects, and more specifically to test whether these two groups differed in the proportion of IgA-producing ASCs with a mucosal tropism in blood, we used an antigen-specific ELISpot assay to measure the number of ASCs producing Spike-specific IgA or IgG after prior enrichment of CD38-expressing cells. Since mucosal ASCs preferentially express tissue specific cell surface homing markers, we also partitioned ASCs expressing integrin β7 which has been proposed to promote the migration of ASCs toward mucosal tissues such as the intestinal mucosae. Human ASCs were enriched from lysed blood using a mixture (1:1) of magnetic beads (Dynabeads Pan Mouse IgG, Invitrogen) coated with monoclonal antibodies (mAbs) to either CD38 (clone HB-7, Biolegend) or β7 integrin (clone FIB504, BD Bioscience) followed by application of a magnetic field. For antigen-specific ASC enumeration, the wells of ELISPOT plates were coated with purified Spike (Sino Biological Europe GmbH) and control antigens (i.e. Bovine Serum Albumin, BSA) as described elsewhere (18). Similarly, immunoglobulin-secreting cells (ISCs) irrespective of the antigen specificity were enumerated in parallel wells coated with a mixture of affinity-purified goat antibodies to human Ig k and λ light chains. After incubation of ASC- and ISC-containing cell suspensions for 3 hours at 37°C, wells were extensively washed with PBS-EDTA and PBS-Tween 20. Next, a mixture of appropriately diluted goat antibodies to human IgA and IgG, respectively labelled with alkaline phosphatase and horseradish peroxidase (Southern Biotech) was added to the wells. Zones of solid phase-bound secreted IgA and IgG antibodies were visualized by stepwise incubation with corresponding enzyme chromogen substrates. After drying, plates were scanned and blue (IgA) and red (IgG) spots enumerated using an automated ELISPOT reader. Total and nominal mucosal ASCs as well as ISCs were enumerated against each SARS-CoV-2 Spike antigen and net ASC and ISC counts were determined after subtracting corresponding non-specific counts detected in control (BSA-coated) wells. Data are expressed as the ASC or ISC number per millilitre of blood.

### Statistical analyses

Data are presented as geometric means and 95% confidence intervals of geometric means with a standard error of the mean (SEM) and medians with interquartile ranges (IQR) for quantitative variables, or as numbers and percentages for categorical variables. Comparison between time points was performed with the Wilcoxon matched-pairs, two-tailed rank test. The Chi-square test was used for comparison of seroconversion rates. Statistical analyses were performed using GraphPad Prism 9·0 (GraphPad Software, Inc., San Diego, CA). Differences were considered significant when the p-value was < 0·05. For a multivariate analysis, a logistic regression using seroconversion as the outcome and risk factors as independent variables was performed. Logistic regressions were conducted in R (R Core Team, 2020).

## Results

We enrolled COVID-19 convalescent PCR-negative patients (n=20) who had been infected with SARS-CoV-2 before the emergence of the delta variant and SARS-CoV-2-naive seronegative subjects (n=23). Subjects in the two groups did not differ in age, gender, Body Mass Index (BMI), and total number of blood leukocytes, lymphocytes and neutrophils (Supplementary Table 1).

Before vaccination, IgA and IgG to Spike and Spike RBD were readily detectable in both NELF (Figure 1) and plasma (Supplementary Figure 2) in COVID-19 convalescent patients. Likewise, NELF (Figure 2) contained antibodies that inhibited the binding of the Wuhan, alpha and delta strains Spike protein to its ACE-2 receptor. A binding inhibition activity was also evidenced in plasma for all the tested variants (Supplementary Figure 3).

**Figure 1:**
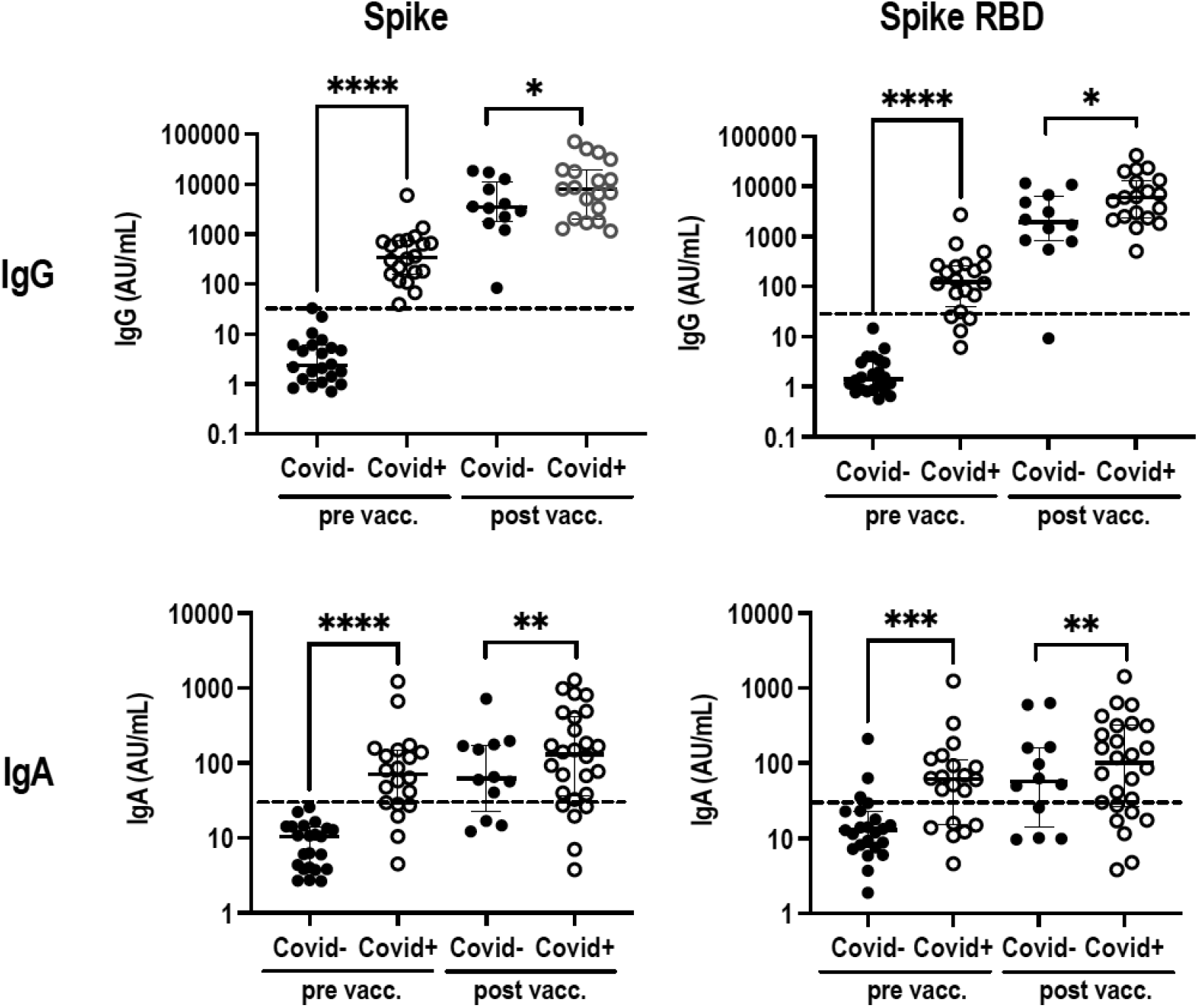
SARS-CoV2-specific IgG and IgA in NELF. NELF from Covid^-^ (filled dots) and Covid^+^ (empty dots) subjects before (pre vacc.) and after (post vacc.) vaccination were analysed for IgG (upper panel) and IgA (lower panel) to Spike and Spike RBD. Data are expressed in arbitrary units per ml (AU/ml) in individual subjects after normalization to the concentration of total IgG and IgA respectively. Medians with the interquartile range are shown. The mean + 2 SD level of IgG and IgA in Covid^-^ subjects before vaccination is indicated by a dotted line. Levels of IgG and IgA to the indicated antigens in Covid^-^ and Covid^+^ subjects before and after vaccination were compared using a two-tailed Wilcoxon-Mann-Whitney test. n.s., not significant; *, p < 0.05; **, p < 0.01; ***, p < 0.001; ****, p < 0.0001.

**Figure 2:**
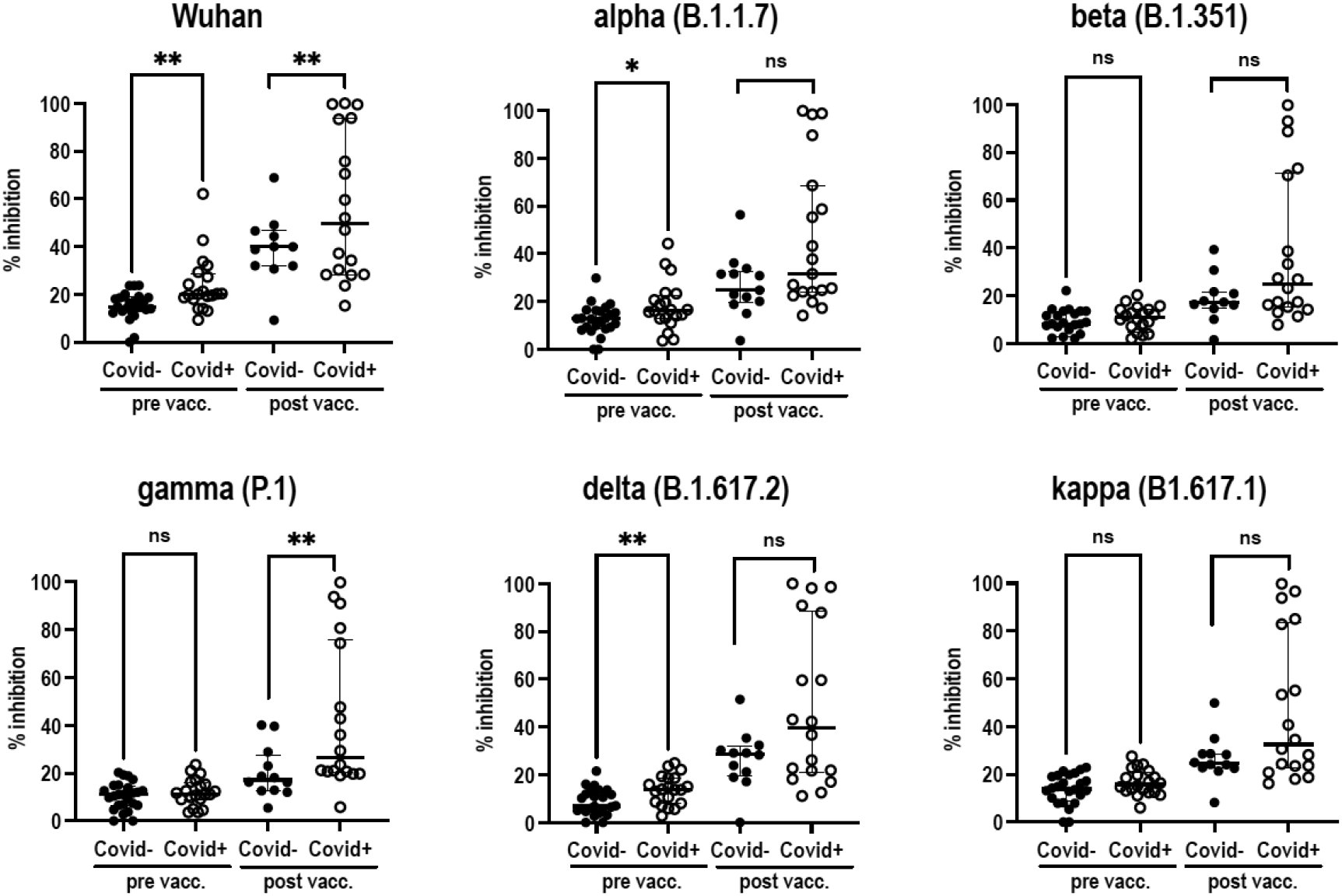
Inhibition of binding of Spike to ACE-2 by NELF antibodies. NELF from Covid^-^ (filled dots) and Covid^+^ (empty dots) subjects before (pre vacc.) and after (post vacc.) vaccination were analysed for the ability to inhibit the binding of the Spike protein of the Wuhan SARS-CoV-2 reference strain and its variants to ACE-2. Percentages of inhibition in individual subjects are shown. The percentage of inhibition of binding in Covid^-^ and Covid^+^ subjects before and after vaccination were compared using a two-tailed Wilcoxon-Mann-Whitney test. n.s., not significant; *, p < 0.05; **, p < 0.01; ***, p < 0.001; ****, p < 0.0001.

We then compared the IgG and IgA mucosal and plasma responses to Spike and Spike RBD in COVID-19 convalescent patients after one vaccine dose and in SARS-CoV-2-naive subjects after two doses. As for NELF, an IgG response to Spike and Spike RBD was observed in all subjects in each group, with these antibodies being 5- to 20-fold more abundant in COVID-19 convalescent patients compared to SARS-CoV-2-naive subjects (Figure 1). In contrast, Spike-specific IgA were only detected in some patients in each group with an increased proportion of responders in COVID-19 convalescent patients compared to SARS-CoV-2-naive subjects (Figure 1). Further, NELF antibodies of COVID-19 convalescent patients were more potent at inhibiting the binding of the Wuhan and gamma variant Spike to ACE-2 compared to those of SARS-CoV-2 naive subjects (Figure 2). As for the plasma, IgG and IgA responses were similar in the two groups both in terms of the proportion of responders (Supplementary Figure 2), IgG and IgA levels among responders (Supplementary Figure 2), and the ability of plasma antibodies to block the binding of Spike to ACE-2 (Supplementary Figure 3).

IgA in NELF are produced by a subpopulation of CD38^+^ ASCs that migrate to the nasal mucosa where they release dimeric IgA. Because the IgA responses in nasal secretions were different in COVID-19 convalescent patients compared to SARS-CoV-2-naive subjects, we hypothesized that these two groups differed in the total number of IgA-producing ASCs in blood, and more specifically in the proportion of IgA-producing ASCs with a mucosal tropism. To test this, we used an antigen-specific ELISpot assay to measure the number of ASCs producing Spike-specific IgA or IgG in blood after prior enrichment of CD38-expressing cells. Because mucosal ASCs preferentially express tissue specific cell surface homing markers, we also partitioned ASCs expressing the integrin β7, which have been proposed to promote the migration of ASCs toward mucosal tissues such as the intestinal mucosae. ASCs producing Spike-specific IgA or IgG were neither detected before vaccination, nor in SARS-CoV-2-naive subjects 7 days after the first vaccine dose (not shown). In contrast, these cells were readily detected after enrichment for both CD38^+^ and β7^+^ cells in both COVID-19 convalescent patients and vaccine-primed SARS-CoV-2-naive subjects after they have received a vaccine dose. The proportion of β7^+^ cells among CD38^+^ was higher in the first group compared to the second one, suggesting an increased proportion of ASCs with a tropism to mucosal tissues (Figure 3).

**Figure 3:**
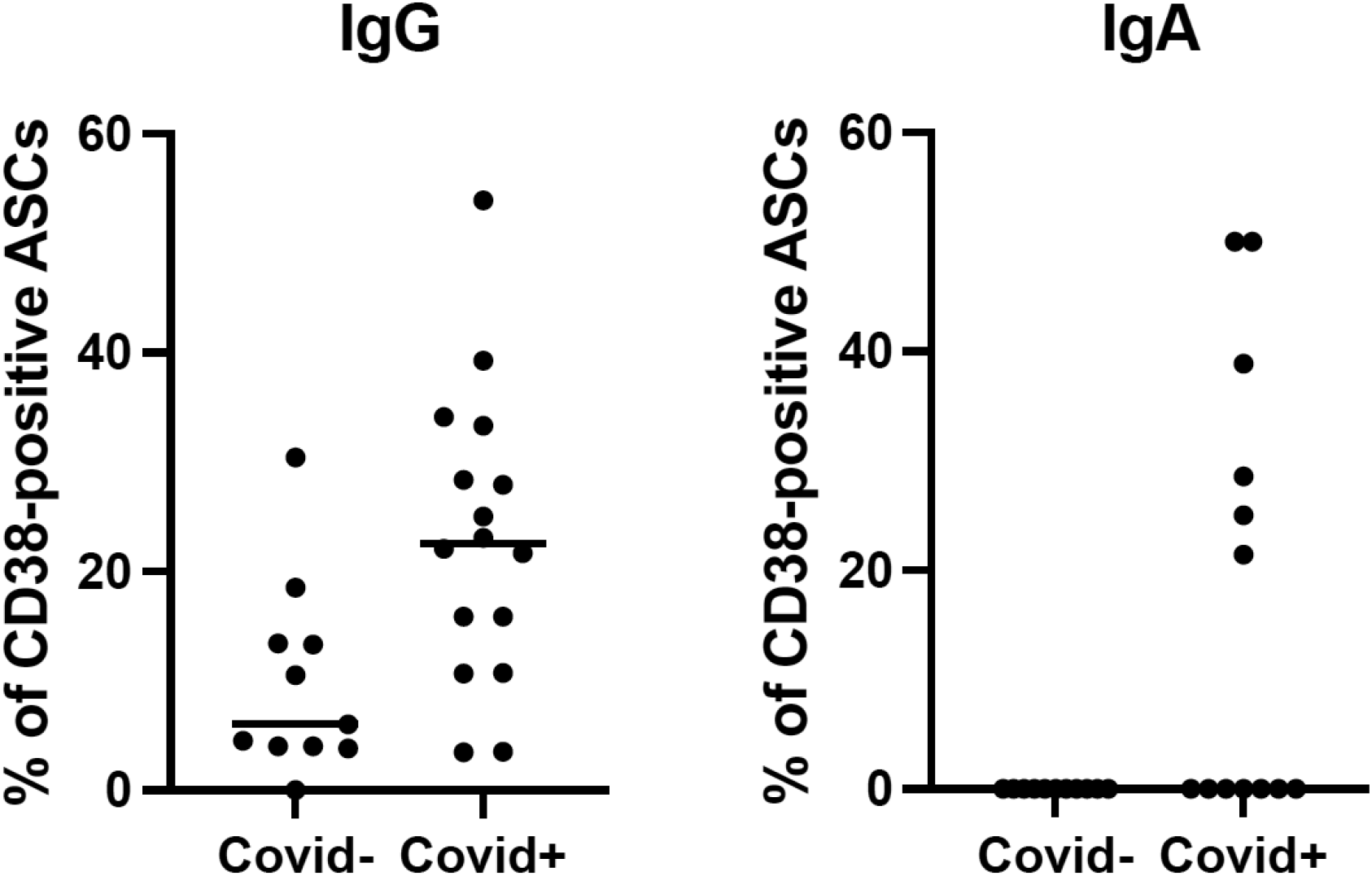
Proportion of ASCs with mucosal tropism. Blood of Covid^-^ subjects taken on day 28 and Covid^+^ subjects on day 7 were analysed for the number of Spike-specific IgA- and IgG-secreting cells in individual subjects after enrichment of CD38^+^ and β7^+^ cells. The proportion of β7^+^ ASCs in the two groups were compared using a two-tailed Wilcoxon-Mann-Whitney test. *, p < 0.05; **, p < 0.01; ***, p < 0.001; ****, p < 0.0001.

## Discussion

In keeping with our initial hypothesis, we found that both the level of Spike-specific IgG and IgA in NELF, the proportion of responders and the ability of NELF antibodies to inhibit the binding of Spike to ACE-2 was higher in COVID-19 convalescent subjects than in vaccine-primed SARS-CoV-2 naive subjects. Further, the proportion of ASCs with a mucosal tropism was higher in COVID-19 convalescent subjects after one vaccine doses compared to SARS-CoV-2 naive subjects after two vaccine doses.

What are the underlying mechanisms that explain this difference? While primary infection with a mucosal pathogen induces the activation of naive B cells in mucosal-associated lymphoid tissue (MALT), parental vaccination induces B cell activation in regional lymph nodes. In both cases, activated B cells proliferate and differentiate into either ASCs or memory B cells. While most ASCs migrate to the bone marrow, homing to other mucosal sites occurs where they produce IgA or IgG, which are eventually transported through epithelial cells by polymeric immunoglobulin receptor (pIgR)- and neonatal Fc receptor (FcRn)-mediated transcytosis, respectively. ASCs homing to selective sites is controlled by the expression of different combinations of adhesion molecules and chemokine receptors, which are induced during initial antigen priming and depends on molecular cues that are governed by the lymph node (LN) microenvironment. For example, ASCs that have been primed in MALTs are instructed to home to mucosal tissues, this mucosal tropism being determined by the expression of chemokine receptors such as CCR9 and CCR20, and integrins such as α4β7 and α4β1. As for memory B cells, they position themselves strategically in secondary lymphoid organs, become tissue-resident at the site of infection or vaccination, or patrol as recirculating cells. During a secondary immune response, memory B cells are induced to differentiate into ASCs the homing tropism of which is determined by the environment in which they have been primed. Based on these findings, we propose the following scenario. Infection with SARS-CoV-2 and parenteral injection of a COVID-19 mRNA vaccine induce the activation of naive Spike-specific B cells and their differentiation into memory B cells in nasal-associated lymphoid tissue (NALT) and regional LN, respectively. In COVID-19 convalescent patients, the injection of a single dose of COVID-19 mRNA vaccine induced memory B cells to differentiate into ASCs with a preferential tropism to mucosa. In contrast, in vaccine-primed SARS-CoV-2-naive subjects, the injection of a second vaccine dose induces B cells to differentiate into ASCs, which are less prone to home to the mucosa. In line with this hypothesis, we found that the proportion of ASCs with mucosal tropism was higher in COVID-19 convalescent patients after the injection of a single vaccine dose compared to SARS-CoV-2-naive subjects after they received two doses.

The inhibition of binding of Spike to ACE-2 by NELF and antibodies in plasma is mediated by RBD-specific IgG and IgA. In keeping with the higher levels of RBD-specific IgG and IgA in COVID-19 convalescent patients compared to vaccine-primed SARS-CoV-2-naive individuals, the inhibition of binding by antibodies in plasma and NELF at baseline was higher in the first group compared to the second one. This was observed for the Wuhan parental strain and its variants including delta. Vaccination further increased inhibition of binding in COVID-19 convalescent patients compared to vaccine-primed subjects. Of note, inhibition of binding by antibodies in plasma was higher in COVID-19 convalescent patients after vaccination compared to SARS-CoV-2 naive subjects after vaccine priming and boosting. While this latter result may appear surprising considering that patients from these two groups exhibited similar amounts of RBD-specific IgG and IgA in plasma, the explanation may lie in the possibly that these antibodies were present in saturating amounts in the assay for inhibition of binding.

The present study has some limitations. Firstly, our study sample consisted of a small cohort of COVID-19 convalescent patients and SARS-CoV-2-naive subjects. Therefore, our results need to be validated with a larger and independent cohort of patients. Further, only 11 Covid^-^ subjects were sampled after the second vaccine dose therefore limiting the statistical power. Secondly, the results showing the impact of vaccination on inhibition of binding by NELF antibodies should be taken with caution because of the high inter-individual variability in the levels of total IgG and IgA in NELF possibly resulting from differences in the production of mucus. Thirdly, COVID-19 convalescent patients were sampled three weeks after vaccination but not at later time points. Therefore, it is possible that the levels of SARS-CoV-2-specific IgA in NELF may have dropped after three weeks. Finally, while mucosal IgA have been demonstrated to prevent virus shedding and transmission in other infectious diseases, this has not been demonstrated in COVID-19 patients or vaccinated SARS-CoV-2 naive subjects.

To our knowledge, this is the first study that documents the induction of mucosal immune responses in the upper airway mucosa, the main site of infection by SARS CoV-2 and as such a major site of person-to-person transmission. Given the exceptionally potent SARS-CoV-2 neutralizing properties of secretory antibodies and particularly secretory IgA (19) the most abundant class of Ig in human secretions, this study indicates that sIgA antibodies should be given special attention as potential correlates or surrogates of immune protection for candidate SARS CoV-2 vaccines.

As a SARS-CoV-2 mucosal vaccine is not yet available, we could not compare directly a mucosal prime-parenteral boost to a parenteral prime-parenteral boost regimen to enhance COVID-19 vaccine immunogenicity. We therefore compared the impact of a parenteral boost in subjects who had been primed by a SARS-CoV-2 infection and in naive individuals for SARS-CoV-2 after parenteral priming. Our results provide a rationale for evaluating a mucosal/systemic prime-boost strategy when COVID-19 mucosal vaccines will be available. Should this vaccination regimen be validated in future trials evaluating efficacy and effectiveness, it may lead to rethinking the practices for vaccination to control infection and transmission of SARS-CoV-2.

Our results may also lead to the establishment of reliable proxy markers of mucosal immunity against SARS-CoV-2 and thereby accelerate licensure of future candidates and SARS-CoV-2 vaccines.

## Data Availability

All data produced in the present study are available upon reasonable request to the authors

## Contributors

CZ, NG and CHM designed the study. NG and CHM wrote the clinical protocol. PH supervised the preparation and storage of biological samples. CHM, CM, SL OG, JBE, JBO, SL, OG and EM, included subjects and collected informed consent. CM and EM collected blood samples and NELF. VT and MA prepared and stored biological samples. EM, TS, PB and CZ performed experiments. ME, VH, TS, AS provided advice. EM and NG analyzed data. NG CM, PH and CZ wrote the manuscript. All authors revised the manuscript and gave final approval.

## Acknowledgments

Research reported in this publication was supported by grants from the Ministère de l’Enseignement Supérieur et de la Recherche, the Conseil Départemental des Alpes Maritimes, the Métropole Nice Côte d’Azur. Special thanks to E. Faidhi, N. Fridlyand, A. Rauscher, E. Maris, the Lauro family and to the many private donators for their generous contribution. The authors thank the subjects who volunteered in this study as well as the medical and paramedical personnel involved in their recruitment and follow-up.

## Data availability statement

The data that support the findings of this study are available from NG.

## Supplementary Materials

**Supplementary Table 1:**
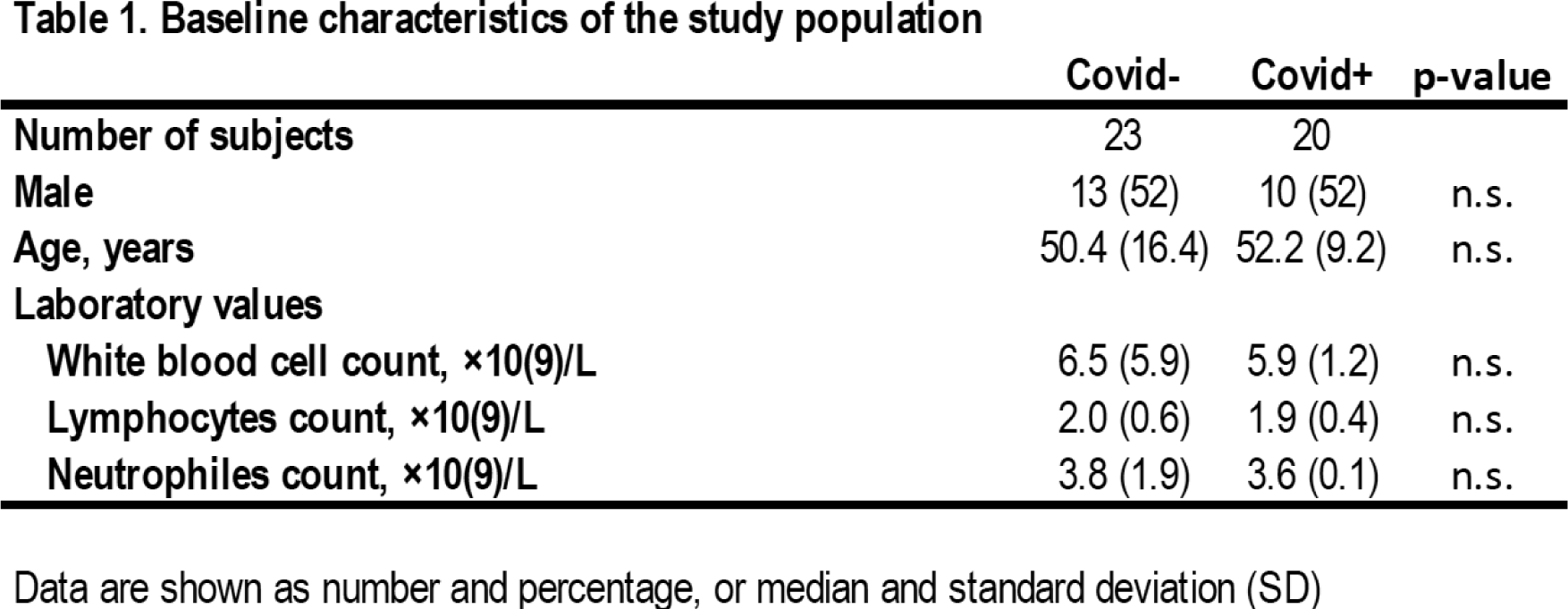
Subjects characteristics: Covid^-^ and Covid^+^ individuals were compared using a Chi-square test for categorical variables and a Mann-Whitney U test for continuous variables respectively and the corresponding p-values were computed. Differences between groups were considered to be statistically significant when corrected p-values were < 0.05. n.s.; not significant.

**Supplementary Figure 1.**
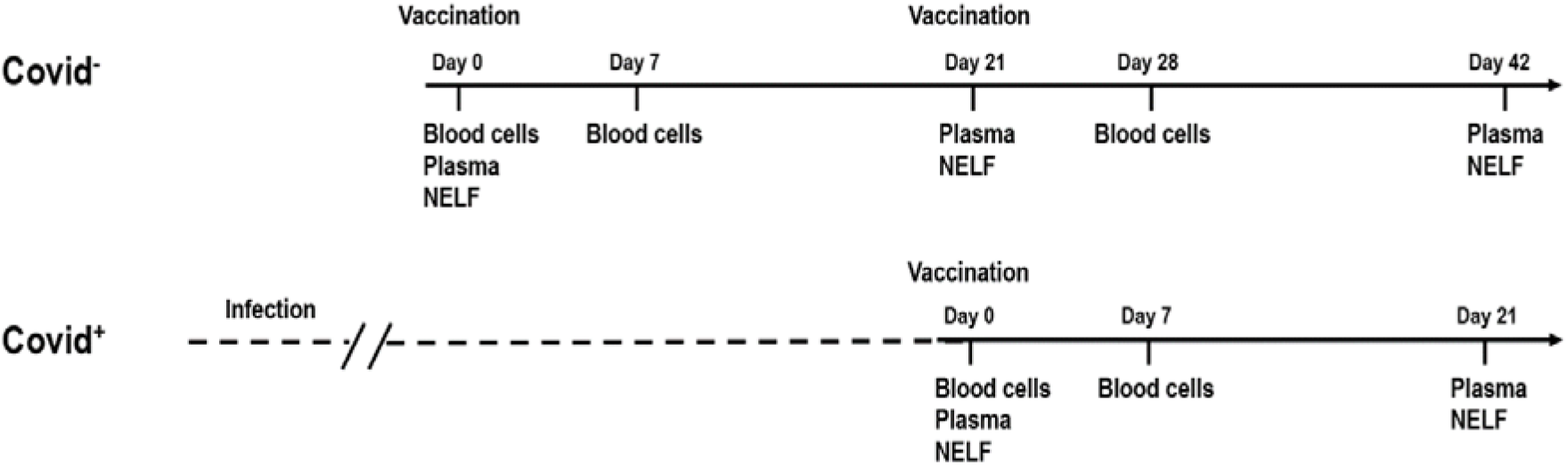
Schematic representation of the clinical protocol: All individuals were included on day 0, vaccinated with the Pfizer mRNA COVID-19 vaccine, and sampled for blood cells on day 0 and day 7 and for plasma and NELF on day 0 and day 21. Covid^-^ individuals were boosted with a second dose of the Pfizer mRNA COVID-19 vaccine on day 21, and sampled for blood cells on day 28 and for plasma and NELF on day 42.

**Supplementary Figure 2.**
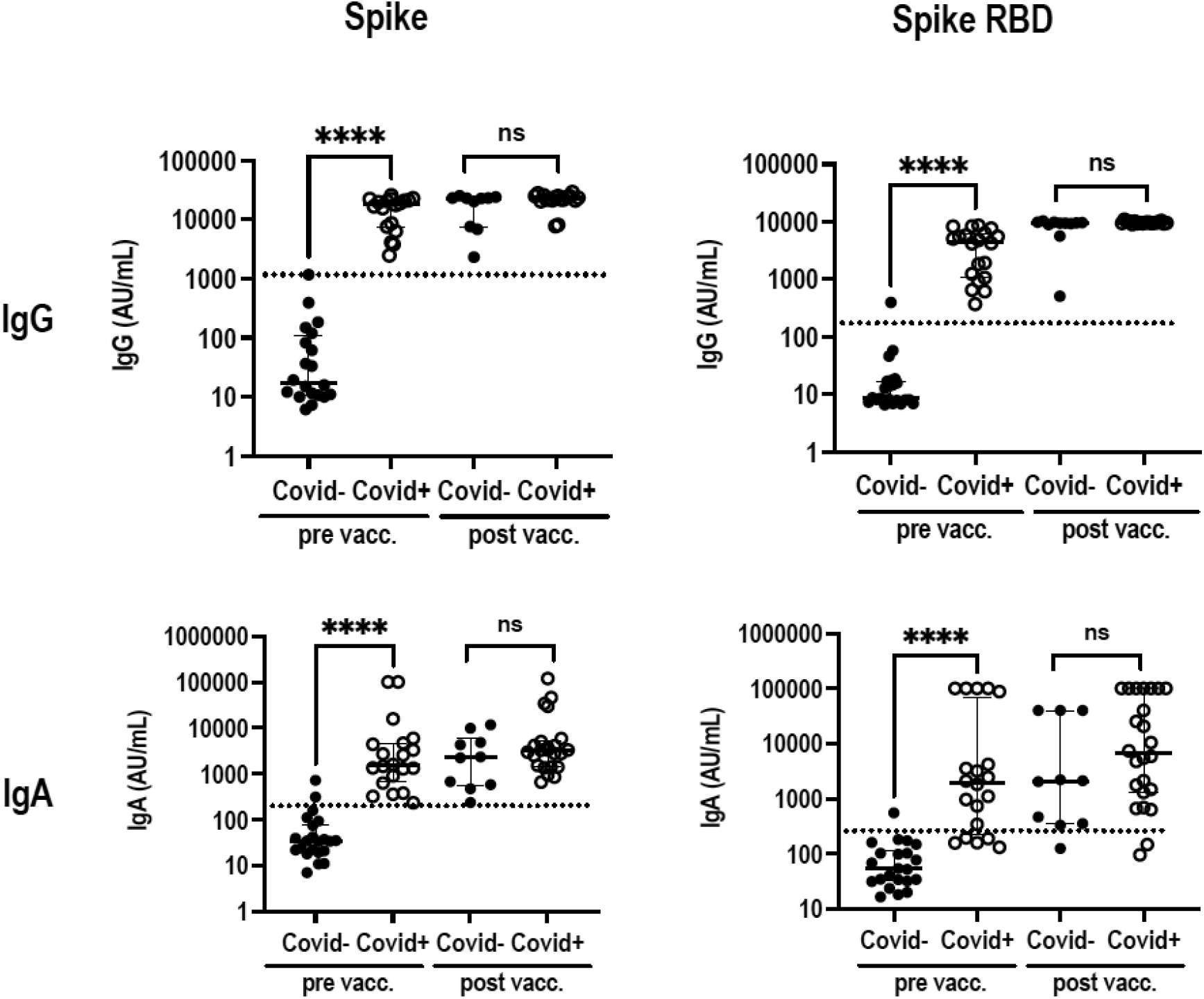
SARS-CoV2-specific IgG and IgA in plasma : Plasma from Covid^-^ (filled dots) and Covid^+^ (empty dots) subjects before (pre vacc.) and after (post vacc.) vaccination were analyzed for IgG (upper panel) and IgA (lower panel) to Spike and Spike RBD. Data are expressed in arbitrary units per ml (AU/ml) in individual subjects after normalization to the concentration of total IgG and IgA respectively. Medians with the interquartile range are shown. The mean + 2 SD level of IgG and IgA in Covid^-^ subjects before vaccination is indicated by a dotted line. Levels of IgG and IgA to the indicated antigens in Covid^-^ and Covid^+^ subjects before and after vaccination were compared using a two-tailed Wilcoxon-Mann-Whitney test. n.s., not significant; *, p < 0.05; **, p < 0.01; ***, p < 0.001; ****, p < 0.0001.

**Supplementary Figure 3.**
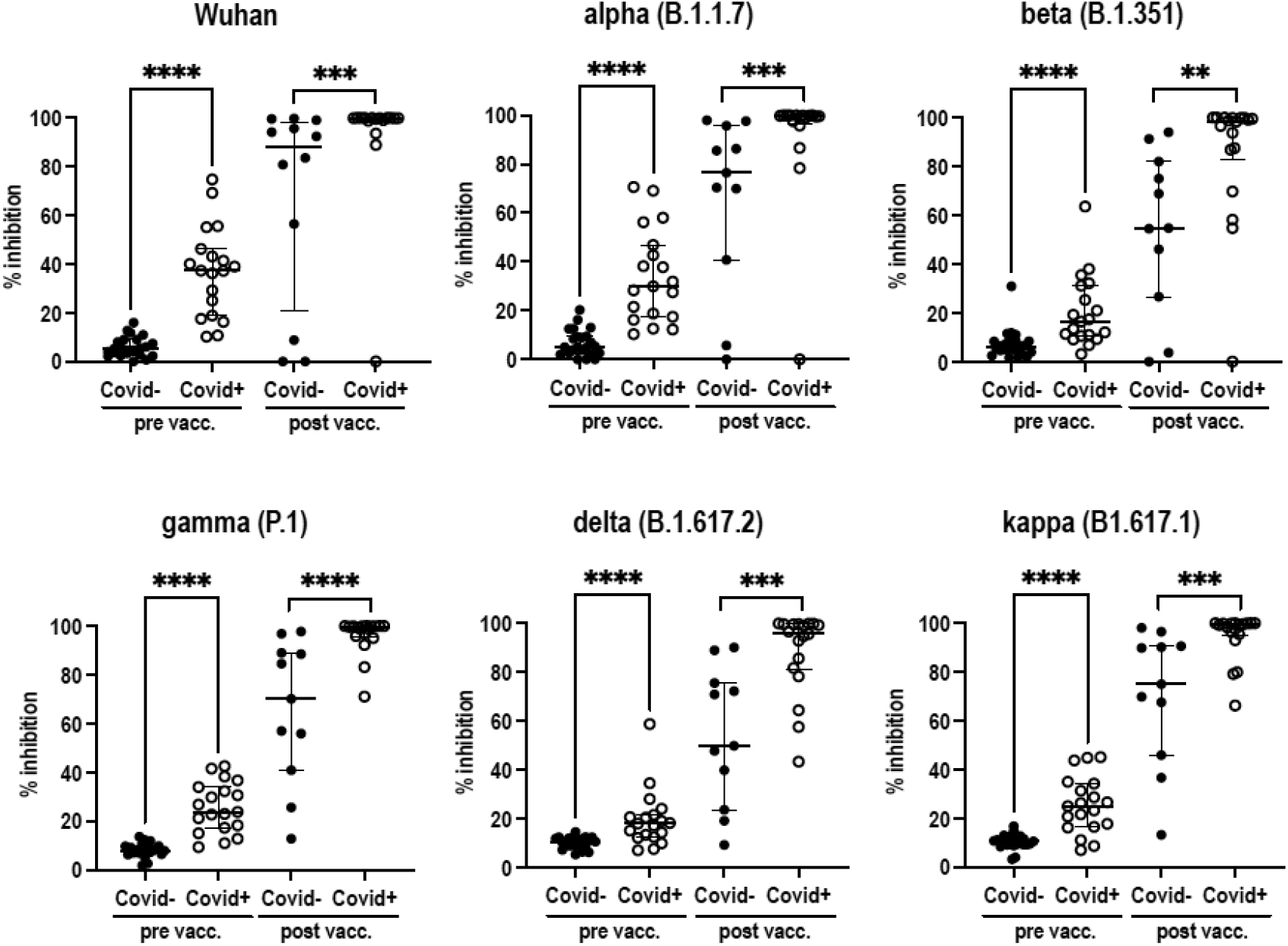
Inhibition of binding of Spike to ACE-2 by plasma antibodies. Plasma from Covid^-^ (filled dots) and Covid^+^ (empty dots) subjects before (pre vacc.) and after (post vacc.) vaccination were analyzed for the ability to inhibit the binding of ACE-2 to the Spike protein of the Wuhan SARS-CoV-2 reference strain and its indicated variants. Percentages of inhibition in individual subjects are shown. The percentage of inhibition of binding in Covid^-^ and Covid^+^ subjects before and after vaccination were compared using a two-tailed Wilcoxon-Mann-Whitney test. n.s., not significant; *, p < 0.05; **, p < 0.01; ***, p < 0.001; ****, p < 0.0001.

## References

1. Lopez Bernal J, Andrews N, Gower C, Gallagher E, Simmons R, Thelwall S, et al. Effectiveness of Covid-19 Vaccines against the B.1.617.2 (Delta) Variant. N Engl J Med. 2021;385(7):585–94.

2. Linsenmeyer K, Gupta K, and Charness ME. Effectiveness of Covid-19 Vaccines against the B.1.617.2 (Delta) Variant. N Engl J Med. 2021.

3. Emani VR, Reddy R, and Goswami S. Effectiveness of Covid-19 Vaccines against the B.1.617.2 (Delta) Variant. N Engl J Med. 2021.

4. Torgovnick J. Effectiveness of Covid-19 Vaccines against the B.1.617.2 (Delta) Variant. N Engl J Med. 2021.

5. Shah ASV, Gribben C, Bishop J, Hanlon P, Caldwell D, Wood R, et al. Effect of Vaccination on Transmission of SARS-CoV-2. N Engl J Med. 2021;385(18):1718–20.

6. Harris RJ, Hall JA, Zaidi A, Andrews NJ, Dunbar JK, and Dabrera G. Effect of Vaccination on Household Transmission of SARS-CoV-2 in England. N Engl J Med. 2021;385(8):759–60.

7. Singanayagam A, Hakki S, Dunning J, Madon KJ, Crone MA, Koycheva A, et al. Community transmission and viral load kinetics of the SARS-CoV-2 delta (B.1.617.2) variant in vaccinated and unvaccinated individuals in the UK: a prospective, longitudinal, cohort study. Lancet Infect Dis. 2021.

8. Begin P, Callum J, Jamula E, Cook R, Heddle NM, Tinmouth A, et al. Convalescent plasma for hospitalized patients with COVID-19: an open-label, randomized controlled trial. Nat Med. 2021.

9. Corthesy B. Multi-faceted functions of secretory IgA at mucosal surfaces. Front Immunol. 2013;4:185.

10. Sterlin D, Mathian A, Miyara M, Mohr A, Anna F, Claer L, et al. IgA dominates the early neutralizing antibody response to SARS-CoV-2. Sci Transl Med. 2021;13(577).

11. Sun L, Kallolimath S, Palt R, Stiasny K, Mayrhofer P, Maresch D, et al. Increased in vitro neutralizing activity of SARS-CoV-2 IgA1 dimers compared to monomers and IgG. Proc Natl Acad Sci U S A. 2021;118(44).

12. Lavelle EC, and Ward RW. Publisher Correction: Mucosal vaccines -fortifying the frontiers. Nat Rev Immunol. 2021.

13. Lavelle EC, and Ward RW. Mucosal vaccines - fortifying the frontiers. Nat Rev Immunol. 2021.

14. Dey A, Molodecky NA, Verma H, Sharma P, Yang JS, Saletti G, et al. Human Circulating Antibody-Producing B Cell as a Predictive Measure of Mucosal Immunity to Poliovirus. PLoS One. 2016;11(1):e0146010.

15. French National Health Authority FNH, HAS ed.; February 2021.

16. Tanga V, Leroy S, Fayada J, Hamila M, Allegra M, Messaoudi Z, et al. Establishment of a Collection of Blood-Derived Products from COVID-19 Patients for Translational Research: Experience of the LPCE Biobank (Nice, France). Biopreserv Biobank. 2020;18(6):517–24.

17. Watelet JB, Gevaert P, Holtappels G, Van Cauwenberge P, and Bachert C. Collection of nasal secretions for immunological analysis. Eur Arch Otorhinolaryngol. 2004;261(5):242–6.

18. Saletti G, Cuburu N, Yang JS, Dey A, and Czerkinsky C. Enzyme-linked immunospot assays for direct ex vivo measurement of vaccine-induced human humoral immune responses in blood. Nat Protoc. 2013;8(6):1073–87.

19. Brandtzaeg P. Secretory IgA: Designed for Anti-Microbial Defense. Front Immunol. 2013;4:222.

